# Bio-cognitive Cut-Points Differentiate Risk *vs.* No Risk for Prodromal Parkinson Cognition in Young Post-mTBI Veterans and Non-mTBI Controls

**DOI:** 10.64898/2026.08.10.26360106

**Authors:** VA Nejtek-Salvatore, R James, G Boehm, H Alphonso, K Brice, Soto, P Braden-Kuhle, K Doshier, MF Salvatore

**Affiliations:** UNT Health of Fort Worth; Texas Christian University; JPS Health Network; Parkinson Foundation; University of Cincinnati; The Parkinson Discovery Institute; Rice University; Vanguard University; Texas Health Resources

**Author notes:** Corresponding Author: Vicki A. Nejtek, Ph.D. 3500 Camp Bowie Blvd. RES 402D Fort Worth, Tx 76107.

**Keywords:** residual mTBI, Parkinson, Veterans, Biomarkers, Early-stage, Prodromal, Young, Non-mTBI

## Abstract

Blood-based (BB) biomarker investigations in Parkinson disease (PD) and in mild traumatic brain injury (mTBI) have substantially grown over the past decade. High risks for PD in young post-mTBI veterans have been inferred from medical record data using actuarial modeling. However, potential utility of BB biomarkers to quantify risks vs. no risk for PD in young post-mTBI veterans has not been established. Previously we reported post-mTBI veterans performed significantly below the standardized normative scores for their age and education level on specific domains of executive functioning, on par with senior aged individuals with early-stage PD. Here, we examined serum brain-derived neurotropic factor (BDNF), ubiquitin C-terminal hydrolase-L1 (UCH-L1), glial fibrillary acidic protein (GFAP), and S100 calcium-binding protein β (S100B) in association with executive functioning outcomes in search of a bio-cognitive model suitable to differentiate risk from no risk for prodromal PD. A reference range of bio-cognitive cut-points were derived from Area Under the Curve (AUC) sensitivity and specificity methods.

Our data revealed two bio-cognitive signatures with reference range cut-points when GFAP was paired with cognitive flexibility / attention scores, and when S100B was paired with categorical / semantic verbal memory scores. Both bio-cognitive signatures revealed prodromal PD risk vs.no-risk parameters that remained relevant for differentiating young veterans who had encountered a past mTBI and those who had not experienced a mTBI. Subjects with early-stage PD who had withstood a mTBI up to 10- to 40-years earlier were also differentiated from those who had no mTBI history. These results indicate the predictive utility of expanding the biomarker field to include reference ranges, cut-points, and specific cognitive domains to estimate risks for PD in a clinic setting. These preliminary data also add value in establishing a quantifiable bio-cognitive risk signature to identify prodromal PD risks in young adults prior to obvious cognitive and motor decline. While encouraging, these data require further follow-up with a larger sample size in a longitudinal design to validate these findings.

## Introduction

Although blood-based biomarker studies in Parkinson disease (PD) and mild traumatic brain injury (mTBI) have substantially grown over the past decade, predicting PD risks in veterans who have experienced a past mTBI remain unclear. In our previous work, we reported that young veterans who had experienced a past mTBI occurring up to 7-years earlier showed cognitive deficits in specific domains that mirrored the results from those in early-stage Parkinson’s disease (PD; Nejtek et al., 2021). Specifically, cognitive flexibility, attention, verbal fluency test scores in veterans with mTBI ranked significantly below the standardized normative scores for their age and education level. These data illustrated premature cognitive decline in veterans with mTBI who performed like senior aged individuals with early-stage PD. In an extension of the parent study, our primary goals were to (1) investigate blood-based, serum biomarkers to biologically characterize the premature cognitive decline we found in the aforementioned cognitive domains; (2) combine serum biomarkers with specific executive domains to identify a bio-cognitive signature that is sensitive and specific enough to discern risks for PD in post-mTBI veterans prior to obvious motor impairment, and (3) determine reference range cut-points for a sensitive bio-cognitive signature to differentiate parameters of risk for PD *vs*. those who are not at risk.

Convincing data suggest that astrocytes play a significant role influencing dopamine (DA) signaling in brain areas impacted by mTBI and PD (Soto et al., 2026; Kasanga et al., 2024; Hier et al, 2021; Schultz et al 2021; Su et al 2012; Lisi et al, 2025; Pittolo et al, 2022; Impellizzeri et al, 2016; Papuc E, Rejdak K., 2020; Haim et al, 2015; Jha et al, 2018; Angelopoulou et al, 2021). In the PD and TBI literature, the astrocytic, calcium-binding peptide signaling protein (S100B) and glial fibrillary acidic protein (GFAP) are elevated in the substantia nigra (SN) and prefrontal cortex (PFC) in rodent PD models *and* in post-mortem PD human tissue compared to controls (Kalia, 2018; Rodriguez et al, 2020; Kasanga et al., 2024; Tang et al, 2023; Angelopoulou et al, 2021; Papuc E, Rejdak K., 2020; Impellizzeri et al, 2016;). Ubiquitin C-terminal hydrolase-L1 (UCH-L1) is released from neurons and help maintain axonal integrity by regulating protein degradation (Wang et al, 2021; Bishop et al, 2016; Ding et al, 2023; Mi and Graham, 2023). Allosteric modulation of astrocytes can inhibit or suppress UCH-L1 in such a way that it loses its ability to protect nigrostriatal neuron viability thereby initiating neuropathology in mTBI and in PD (Wang et al, 2021; Bishop et al, 2016; Ding et al, 2023; Mi and Graham, 2023; Largares et al, 2023; Ng et al, 2020). Moreover, data suggest that peripheral serum levels of UCH-L1, GFAP, and S100B may be viable biomarkers of PD and TBI severity (Largares et al., 2024) and cognitive decline (Bishop et al, 2016; Mi and Graham, 2023; Tang et al, 2023; Schultz et al., 2023) that suggest these biomarkers may predict risks for PD in post-mTBI individuals.

Brain-derived neurotropic factor (BDNF) and glial cell-derived neurotrophic factor (GDNF) are well-studied in neurodegenerative research. Some studies suggest that early-stage *de novo* PD patients have significantly *lower* levels of serum BDNF (Hernandez-Vara et al, 2020; Khalil et al, 2016; Baquet et al, 2005). Lower BDNF levels are associated with dysregulation of nigrostriatal mechanisms and lower global cognition outcomes when compared to healthy controls (Khalil et al. 2016; Wang et al., 2016). BDNF may protect substantia nigra neurons from atrophy and dysfunction (Nagahara et al, 2009; Zuccato and Cattaneo, 2009; Baquet et al, 2005), although our recent work suggests little involvement of BDNF expression to protect against nigrostriatal neuron loss (Kasanga et al., 2023).

GDNF levels have been shown to *<u>increase</u>* in response to astrocytic dysfunction which can also increase DA signaling in nigrostriatal neurons (Salvatore 2004; Kasanga et al, 2023; Kasanga et al, 2019). Moreover, our preclinical work shows that the GDNF family receptor (GFR) GFR-α1, progressively decreases in striatum and SN – both of which are key brain areas associated with PD (Kasanga et al., 2019; 2023). Thus, whether neurotropic factors are sufficiently sensitive and specific enough to predict prodromal or early-stage PD from a remote mTBI is unclear (Hier et al, 2021).

The bulk of TBI data are derived from various protocols and diagnostic methodologies, varied inclusion / exclusion criteria, and heterogenous TBI severities that would be impossible to link to PD risks (Riley, 2020; Fiandaca et al.,2018; Huie et al., 2021; Whitehouse et al., 2025). For PD studies, the biomarker field has been flooded with hundreds of proteins obtained from cerebral spinal fluid, plasma, serum, and the metabolome which results in a confounding mix of specimen sources (Fiandaca et al.,2018; Huie et al, 2019; Hier et al., 2021; Whitehouse et al., 2025; Cui et al, 2022; Kocheril et al, 2022). Thus, it was important to discern if the premature cognitive decline we found in our parent study (Nejtek 2021) could be strengthened with serum biomarkers in search of a bio-cognitive signature of post-mTBI risks to add to the list of prodromal PD symptoms (Berg and Postuma, 2018). Here, we achieved our goals and discovered not one but two bio-cognitive signatures to help us identify ‘yes vs. no’ risk for PD or some other neurodegenerative disorder (oND).

## Methods and Materials

### Study Design and Participants

As a follow-up to the parent study (Nejtek et al, 2021), a prospective, cross-sectional, proof-of-concept, matched-control study was designed to explore serum biomarkers associated with the cognitive outcomes previously reported among veterans with (+) and without (-) mTBI, age and IQ-matched controls, and early-stage non-demented PD subjects (Nejtek et al., 2021). Institutional Review Board (IRB) approval was obtained through the North Texas Regional IRB (http://www.irb.net.org). Written informed consent was obtained from all volunteers prior to study enrollment. A community-based participatory research approach was used to recruit all subjects. Although the complete study protocol is reported in detail elsewhere (Nejtek et al., 2021), we briefly highlight the following study procedures.

### Inclusion / Exclusion Criteria, Neurological and Mood State

Inclusion eligibility for veterans (+) mTBI were those incurring a non-penetrating, combat-related mTBI ≤ 7-years of study enrollment whose TBI grade was verified as mild using Veteran Administration Department of Defense consensus criteria (Fortier et al, 2014; Corrigan et al., 2007; Schwab et al, 2006). Inclusion of PD subjects were those in the early-stage of the disease categorized as Hoehn & Yahr 1.0 - 1.5 (Hoehn & Yahr, 1967; Goetz et al., 2004), the Movement Disorder Society Task Force on Rating Scales for Parkinson’s Disease (Shulman et al, 2009), and each patient’s treating neurologist reports. Individuals with Parkinson disease were excluded if they had any other unremitted co-occurring central nervous system disorder or condition (i.e. stroke, chronic migraines, epilepsy, seizures, etc.) that could introduce bias or prevent completion of study procedures.

Cumulative scores from the Posttraumatic-Stress-Disorder Checklist (PTSD-C) were used to exclude veterans scoring higher than the 50-point threshold for current PTSD in military personnel, and higher than the 44-point threshold for healthy civilian controls (Weathers et al., 1993). The Geriatric Depression Scale (GDS) excluded all subjects scoring higher than the ‘normal’ range (0-9) (Yesavage et al., 1983). A minimum score of 24 on the Mini Mental State Exam (MMSE) excluded subjects with frank global dementia or mild cognitive impairment (Crum et al, 1993; Folstein et al., 1975). The Mini Neuropsychiatric Interview (MINI; version 6.0) excluded subjects with currently unremitted mood, psychotic, or substance use disorders (also verified with urine drug screens) and subjects experiencing active suicidal thoughts [Sheehan et al, 1998].

### Cognitive Assessments

Cognitive assessments from the National Institute of Neurological Disorders and Stroke Common Data Elements list of well-established cognitive tests with validated normative scores and percentiles stratified by age and education were used (Tombaugh et al, 2004; Lezak et al, 2004; Bowie and Harvey, 2006; Ashendorf et al., 2008; Tombaugh et al, 1999). Previously, veterans + mTBI ranked in the 20^th^ percentile (m=33.5 +/-11.3; low average) on the TMT-A and ranked in the 10^th^ percentile (m=78.3 +/- 29.8) on the TMT-B indicating below average cognitive flexibility and strategic decision-making on both tests (Nejtek et al, 2021). Performance of senior subjects with early-stage PD also showed below average cognitive flexibility and decision-making ranking <35^th^ percentile (m=36.8 +/- 11.8) on TMT-A and ranking <10^th^ percentile (m=98.1 +/- 56.2) on TMT-B (Tombaugh et al, 2004; Lezak et al, 2004; Bowie and Harvey, 2006; Ashendorf et al., 2008; Tombaugh et al, 1999). Thus, TMT-A and B scores were the primary cognitive outcomes to link to biological markers to characterize a bio-cognitive profile associated with mTBI-related risks for early-stage PD. Secondary cognitive outcomes were total number of correct words produced in 60 seconds on Controlled Oral Word Association Tests F.A.S. (i.e. phonetic memory) and Animal Naming (i.e. semantic memory) (Tombaugh et al, 1992; Lezak et al, 2004; Strauss et al, 2006; for more detail, please see Nejtek et al, 2021).

### Blood Collection and Processing

Between the hours of 10:00 a.m. and 3:00p.m., all subjects provided a minimum 4-hour *<u>fasting</u>* blood sample collected from the median cubital vein using a 21- to 23-gauge butterfly needle depending on each subject’s needs. At least two 10mL vacutainer (serum separator) tubes were filled and allowed to clot for a minimum of 30-min at room temperature in a vertical position (Fisher Scientific). Samples were centrifuged within 1-hr of collection for a minimum 10-min at 2500 x *g* to obtain serum. Next, 1.0 mL aliquots of serum were pipetted into cryovial tubes and placed in a -80° C freezer within 2-hrs of collection for storage until specimens could be assayed.

### Biomarker Assay Methods

All laboratory assay staff (GB, KB, PB) were blind to the independent group identification of all samples. All biomarkers were assayed in duplicate for quality assurance. BDNF and GDNF were assayed using the SECTOR Imager 2400A from Meso Scale Discovery (MSD, Rockville, MD) employing a multiplex biomarker platform with high-sensitivity anti-body-based electrochemiluminescence (ECL) kits. Plates were read on an MESO QuickPlex SQ 120 Instrument. Serum samples for BDNF were diluted 1:32 prior to assay. The MSD platform is extensively used to assay biomarkers associated with a range of human disease. ECL measures have well-established sensitivity indices that require less volume than conventional enzyme-linked immunosorbent assays (ELISA). S100B, UCHL-1, and GFAP samples were assayed with ELISA (Biomatik, Wilmington, DE) and were determined using a BMG LabTech FLUOstar Omega plate reader (BMG LabTech, Cary, NC) at a wavelength of 450 nm. Serum was run neat for these analytes. All serum levels are reported as *pg/mL*.

### Statistical Analyses

A General Linear Model (GLM) MANOVA was used to compare unequal sample groups on biomarker concentration levels (*pg/mL*). Biomarkers were then examined in combination with cognitive test results to find a bio-cognitive risk profile characterizing mTBI-related PD risks. Games-Howell post-hoc analyses were used to correct for multiple comparisons, assuming unequal group variance. Next, to identify biomarker and cognitive relationships, a Spearman rank correlation was used. Any significant biomarker-cognitive relationships revealed were further analyzed using a partial correlation to control for *lifetime* years since mTBI was experienced.

First, the partial correlation results guided our decision to aggregate all group data veterans + mTBI plus those who had ever experienced an mTBI in their lifetime (remote past), as a ‘YES risk’ for PD, and those who had *<u>never</u>* in their lifetime experienced any type of head injury as a ‘NO risk’. Second, we conducted a nonparametric Mann-Whitney U test (without assuming normality) to identify the most clinically relevant bio-cognitive marker of mTBI-related PD risks (YES risk *vs*. NO risk). The nonparametric condition allows all possible values to be paired and analyzed without specifying the assumptions or the distribution of the outcomes. Third, Receiver Operating Characteristic (ROC) and nonparametric Area Under the Curve (AUC) methods were applied to discriminate between the YES vs. NO risk groups in search of accurate sensitivity (true positive rate = probability of accurate detection of disease or disease risk *<u>is present</u>*) and specificity (false positive rate = probability that the disease or risk of disease is accurately *<u>not present</u>*) of bio-cognitive parameters that would best predict mTBI-related PD ‘risks’ vs. ‘no risk’ (Fan et al.2006; Hassanzad and Hajian-Tilaki, 2024, Hajian-Tilaki, 2013; Simundic, 2009).

Cognitive tests are already standardized, and norms are well-established according to age and education. Cognitive raw scores were used compared against standardized norms for ‘YES’ and ‘NO’ risk groups and cut-points with reference ranges were analyzed alongside serum biomarkers. As parameters for serum biomarkers for post-mTBI risk groups are currently unknown, we used Classifier Evaluation Metrics with Youden’s cut-offs to help identify a reference range with the most sensitive and specific parameters that could reasonably predict or at least screen for mTBI-related risks for PD (Hassanzad and Hajian-Tilaki, 2024;Hajian-Tilaki, 2013; Simundic, 2009; Ruopp et al, 2008). As log transformations cannot produce normal distributions and can actually induce errors (West, 2022), raw data were used to determine biomarker levels (*pg/mL*) + cognitive test scores to create a bio-cognitive profile. A 95% confidence interval and alpha 0.05 were used to signify statistical significance (SPSS, version 30, IBM Corp., Armonk, N.Y., USA).

## Results

### Demographics

As determined in the parent study (Nejtek et al., 2021), there were 27 Veterans (+) mTBI, age- and IQ-matched veterans (-) mTBI (referred henceforth as simply ‘veterans’) N=27, 30 healthy matched controls (*x̄ = 32.4 +/- 4.8 years old*), and 27 early-stage PD subjects (*x̄ = 68.5 +/- 8.2 years old*). Although the entire sample consisted of more men (N=79) than women (N=35), there were no significant sex differences dispersed among the 4 groups *(X^2^ (3, N=114) = 3.69, p=.30)*, nor significant group differences in IQ (*F(3,110) = 1.92, p = .13*). Similarly, race/ethnicity dispersion was not significantly different among the 4 groups, [Non-Hispanic Caucasian / European (N=70), Hispanic / Latinx / Mexican American (N=29), or Black / African American (N=15) origins *[X^2^ (6, N=114) = 8.75, p=.18*)].

### mTBI

The mean number of years since an mTBI event in veterans was 5.09+/- 1.9, and the mean number of mTBI events during military service was 3.6 +/- 2.8. Event-related mTBI events reported were blasts/explosions (n = 9), motorized accidents (n =7), physical assaults (n = 6), or falls (n = 5) occurring in Iraq, Afghanistan, Persian Gulf, Saudi Arabia, Haiti, South Korea, Asia, and South America. Frequency of remote mTBI occurring > 10-years ago were operationally defined as ‘remote’ among the other groups as shown in Figure 1.

**Figure 1.**
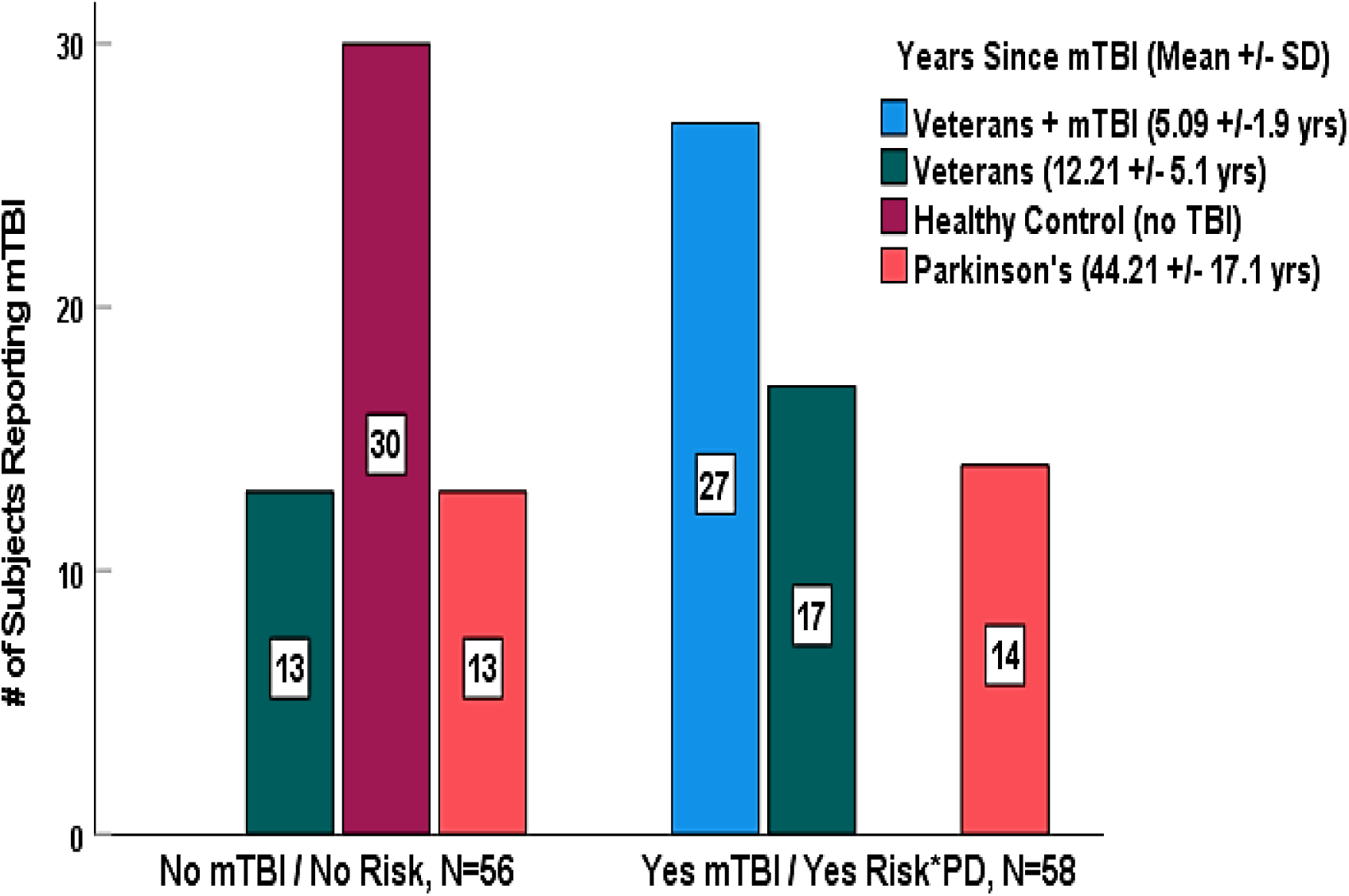
YES or NO mTBI Lifetime Events Per Group (Mean Years Since an mTBI Event)

### Clinical Characteristics

Recall that subjects were excluded if the MMSE indicated global cognitive impairment or frank dementia. Subjects were not clinically depressed (GDS); they did not qualify for a current PTSD diagnosis; and neither outcome influenced any cognitive test scores. (see Nejtek et al., 2021), Due to inter-independent variation of complex dosing regimens, levodopa daily dose equivalents were calculated to determine the PD group mean dose (*630.3, C.I. 445.9 – 814.7*). Subjects with PD receiving levodopa therapy were assessed in the “ON” medication state. There were two de novo subjects. There was no influence of medication on any cognitive test scores (Nejtek et al., 2021).

### Biomarker LOD & CV

Of all the biomarkers analyzed, ∼68% of the GDNF samples fell below the limit of detection (LOD) of the lowest standard curve value. For quality assurance, GDNF samples were re-run with highly sensitive ELISA kits from a different vendor (R&D Systems, Minneapolis, MN). Again, we were unable to detect a reliable GDNF LOD value. With further data verification, we found that standard deviations for GDNF levels among each group varied from 59% to 82% resulting in an overall 55% above the mean for the entire sample suggesting unreliability. Thus, no further analysis of GDNF was conducted.

The intra- and inter-assay coefficients of variance (CVs) for the remaining analytes were all within acceptable ranges: (A) 8.52 and 7.04, for S100B, (B) 5.04 and 9.78 for UCHL-1, (C) 3.12 and 8.46 for BDNF, and (D) 7.26 and 2.61 for GFAP. There were no medication effects on BDNF (*F(1,23) = .07, p = .79),* GFAP *(F(1,23) = .51, p = .48),* UCH-L1*(F(1,23) = .32, p = .58),* or S100B *(F(1,23) =1.02, p = .32*).

### Biomarker * Experimental Group Comparisons

Table 1 shows serum biomarker group comparisons and post hoc analyses. BDNF levels were significantly lower in veterans + mTBI compared to PD subjects (*p = .03*), veteran controls (*p = .05*), and healthy controls (*p =.02*). The PD group showed similar BDNF levels as the young healthy control group (*p = 1.0*) and the young veteran control group (*p = .99*). These similarities may be due to the early stage of PD with fewer negative symptoms than in more severe states, or active lifestyle factors may have played a role since there were no medication effects. In contrast, there were no statistically significant differences in GFAP levels between PD subjects and veterans + mTBI (*p=.29*). In fact, both PD and veteran +mTBI groups had higher GFAP (*pg/mL*) levels than any other group. Neither UCH-L1 nor S100B levels were significantly different among any of the groups, although veterans +mTBI had the lowest UCH-L1 levels and highest S100B levels than any other group.

**Table 1.** Serum Biomarkers Group Comparisons.

| Serum Biomarker x Group |  | Statistics |  |  | Post hoc Analyses |  |  |  |
| --- | --- | --- | --- | --- | --- | --- | --- | --- |
| **BDNF [F (3,100)=4.08, p>.009]** | **M** | **STD** | **95% C.I.** |  | **group** | **p value** | **group** | **p value** |
| a. veterans + mTBI | 9297.09 | 3673.08 | 7746.08 | 10848.10 | *ad* | 0.03 | *ab* | 0.05 |
| b. veterans | 11948.39 | 3476.48 | 10600.35 | 13296.43 | *bd* | 0.99 | *bc* | 0.99 |
| c. healthy controls | 12260.12 | 3602.40 | 10914.97 | 13605.28 | *cd* | 1.00 | *ac* | 0.02 |
| d. Parkinson disease | 12179.04 | 3131.77 | 10790.49 | 13567.59 |  |  |  |  |
| **GFAP [F (3,105)=6.24, p>0.005]** | **M** | **STD** | **95% C.I.** |  | **group** | **p value** | **group** | **p value** |
| a. veterans + mTBI | 51.97 | 62.34 | 26.79 | 77.15 | *ad* | 0.29 | *ab* | 0.56 |
| b. veterans | 34.57 | 28.27 | 23.82 | 45.32 | *bd* | 0.03 | *bc* | 0.85 |
| c. healthy controls | 29.66 | 16.21 | 23.38 | 35.95 | *cd* | 0.02 | *ac* | 0.31 |
| d. Parkinson disease | 93.59 | 100.65 | 52.94 | 134.25 |  |  |  |  |
| **UCH-L1 [F(3,101) = 1.54, p=0.21]** | **M** | **STD** | **95% C.I.** |  | **group** | **p value** | **group** | **p value** |
| a. veterans + mTBI | 665.34 | 452.77 | 469.55 | 861.13 | *ad* | 0.20 | *ab* | 0.24 |
| b. veterans | 972.14 | 674.05 | 705.50 | 1238.78 | *bd* | 1.00 | *bc* | 0.99 |
| c. healthy controls | 1039.56 | 827.83 | 730.44 | 1348.67 | *cd* | 0.99 | *ac* | 0.17 |
| d. Parkinson disease | 973.66 | 605.22 | 723.84 | 1223.48 |  |  |  |  |
| **S100B [F(3,101) = .148, p=0.93]** | **M** | **STD** | **95% C.I.** |  | **group** | **p value** | **group** | **p value** |
| a. veterans + mTBI | 439.45 | 587.50 | 202.15 | 676.75 | *ad* | 0.99 | *ab* | 0.96 |
| b. veterans | 376.11 | 297.62 | 250.43 | 501.78 | *bd* | 0.98 | *bc* | 0.85 |
| c. healthy controls | 438.27 | 247.91 | 342.15 | 534.40 | *cd* | 0.99 | *ac* | 1.00 |
| d. Parkinson disease | 411.95 | 328.31 | 282.08 | 541.83 |  |  |  |  |

### Biomarker and Cognitive Relationships According to mTBI Risks for PD

The Spearman correlation revealed significant relationships between GFAP and cognitive flexibility test scores as measured by TMT-A [*r(114) = .230, p < .014*] and TMT-B [*r(114) = .350, p < .0001*]. Non-significant trends between UCH-L1 and FAS [*r(113) = .177, p=.061],* and S100B and Animal Naming [*r(113) = .170, p=.072]* were noted. There were no significant relationships nor trends between BDNF and any cognitive outcome. As identifying a bio-cognitive marker was integral to fulfill the primary study goal, no further analyses with BDNF were conducted.

To determine if there were any remote *lifetime* mTBIs in the entire sample that could have potentially influenced bio-cognitive relationships, we collapsed the groups into ‘YES’ *vs* ‘NO’ lifetime mTBIs. Figure 1 shows the aggregated number of individuals who reported *<u>ever</u>* experiencing a remote (> 10-yrs. ago) mTBI in their lifetime, veterans +mTBI, and those who reported *<u>never</u>* experiencing mTBI (e.g. YES *vs.* NO group). The ‘YES’ risk group included 52% of PD subjects, 57% veteran controls, and 100% of the veterans + mTBI. The ‘NO’ risk group included 100% of healthy controls, 48% of PD subjects, and 43% of veteran controls. A partial correlation to control for lifetime mTBI events was then used to reexamine biomarker-cognitive relationships. Statistically controlling for lifetime mTBI events abolished all significant bio-cognitive relationships we reported previously. This suggests that a mTBI occurring at any time during a lifetime may initiate subtle residual effects that are likely unrecognized by clinicians as impairments are present only in specific cognitive domains.

### Nonparametric Comparisons YES vs. NO mTBI Lifetime Risk Event Group

As collapsing the 4-group sample into two ‘YES’ ‘NO’ groups could have created a skewed distribution, a nonparametric Mann-Whitney U test was used to analyze the mean ranks for each marker. Table 2 shows significant differences between YES *vs.* NO groups with GFAP biomarker levels, TMT-A and TMT-B cognitive scores significantly higher in the ‘YES’ risk group. Higher scores on TMT-A and TMT-B suggest that poorer attention, cognitive *<u>in</u>*flexibility, and worse strategic decision-making is associated with higher serum GFAP thereby creating a viable bio-cognitive GFAP+TMT-A+TMT-B profile marker. Likewise, the ‘YES’ risk group also showed significantly lower S100B levels and fewer animals named in 60-seconds (Animal Naming task) suggesting poorer categorical / semantic memory and verbal fluency than the ‘NO’ risk group associated with lower S100B. Consequently, these two bio-cognitive profiles were entered into ROC / AUC model to identify specificity and sensitivity parameters to reveal a range of cut-points to infer mTBI-related PD risks. As the bio-cognitive profile of UCH-L1 and FAS did not significantly differ between groups, this bio-cognitive profile was not further evaluated.

**Table 2.** Nonparametric Comparisons of YES vs. NO mTBI Parkinson’s Risk Groups.

| **Table 2** |  |  |  |  |  |  |  |
| --- | --- | --- | --- | --- | --- | --- | --- |
| **Bio-Cognitive Markers** | **Risk** | **N** | **Wilcoxon W** | **Z-value** | **Mann-Whitney U** | **Asymp. Sig.** | **Monte Carlo Sig.** |
| UCH-L1 | No Risk | 54 | 2551.0 | -0.16 | 1331 | 0.533 | 0.534 |
|  | Yes Risk | 53 |  |  |  |  |  |
| S100B | No Risk | 51 | 2304.0 | -2.64 | 926 | **0.008** | **0.007** |
|  | Yes Risk | 52 |  |  |  |  |  |
| GFAP | No Risk | 51 | 2257.0 | -2.34 | 931 | **0.019** | **0.016** |
|  | Yes Risk | 50 |  |  |  |  |  |
| TMT-A | No Risk | 56 | 2790.5 | -2.43 | 1194.5 | **0.015** | **0.014** |
|  | Yes Risk | 58 |  |  |  |  |  |
| TMT-B | No Risk | 56 | 2522.5 | -3.95 | 926.5 | **0.000** | **0.000** |
|  | Yes Risk | 58 |  |  |  |  |  |
| ANIMAL NAMING | No Risk | 56 | 2849.5 | -2.76 | 1138.5 | **0.006** | **0.007** |
|  | Yes Risk | 58 |  |  |  |  |  |
| FAS | No Risk | 56 | 3191 | -0.16 | 1595 | 0.869 | 0.881 |
|  | Yes Risk | 58 |  |  |  |  |  |

In this proof-of-concept study, we anticipated that sensitivity and specificity might be approximately equal in clinical relevance for predicting probability cut-offs. Therefore, Classifier Evaluation Metrics (CEM) to obtain maximum Kolmogorov-Smirnov and Youden’s cut-off values were also used as suggested in the literature (Hassanzad et al, 2024; Haijan-Tilaki, 2013; Šimundić, 2009; Ruopp et al., 2008; Fan et al., 2006).

### Parkinson Disease Risk Identification Using ROC and AUC Analyses for Diagnostic Accuracy

Table 3 shows the AUC for the two bio-cognitive profiles deemed most appropriate to determine clinical relevance with low to moderately high AUC. Figure 2 illustrates the ROC for the GFAP + TMT-A + TMT-B bio-cognitive profile predicting a moderately high risk for PD with very good diagnostic accuracy (*AUC 0.83, 0.75, and 0.81, respectively*) where the reference line represents chance. Coordinates of the ROC curve show 100% sensitivity and 43% specificity for GFAP at ∼30.0 *pg/mL.* Youden’s cut-off for GFAP was 39.83 *pg/mL* (maximum Kolmogorov-Smirnov (K-S) of .63) with coordinates of the ROC curve of 93% sensitivity and 30% specificity. A GFAP level **>30** *pg/mL* is a cut-off that has excellent diagnostic reliability at 100% sensitivity followed by **>39.83** *pg/mL* at 93% sensitivity that might be used as a cut-off range indicating a clinically relevant bio-cognitive risk for PD or some other neurodegenerative disorder (oND). In our sample, GFAP specificity coordinates for PD ranged from 50% for **29.2** to 75% for **15.8** *pg/mL* to 93% for **10.83** *pg/mL* with sensitivity remaining at 100%. Hence, the lower the GFAP the less risk for PD/oND.

**Figure 2.**
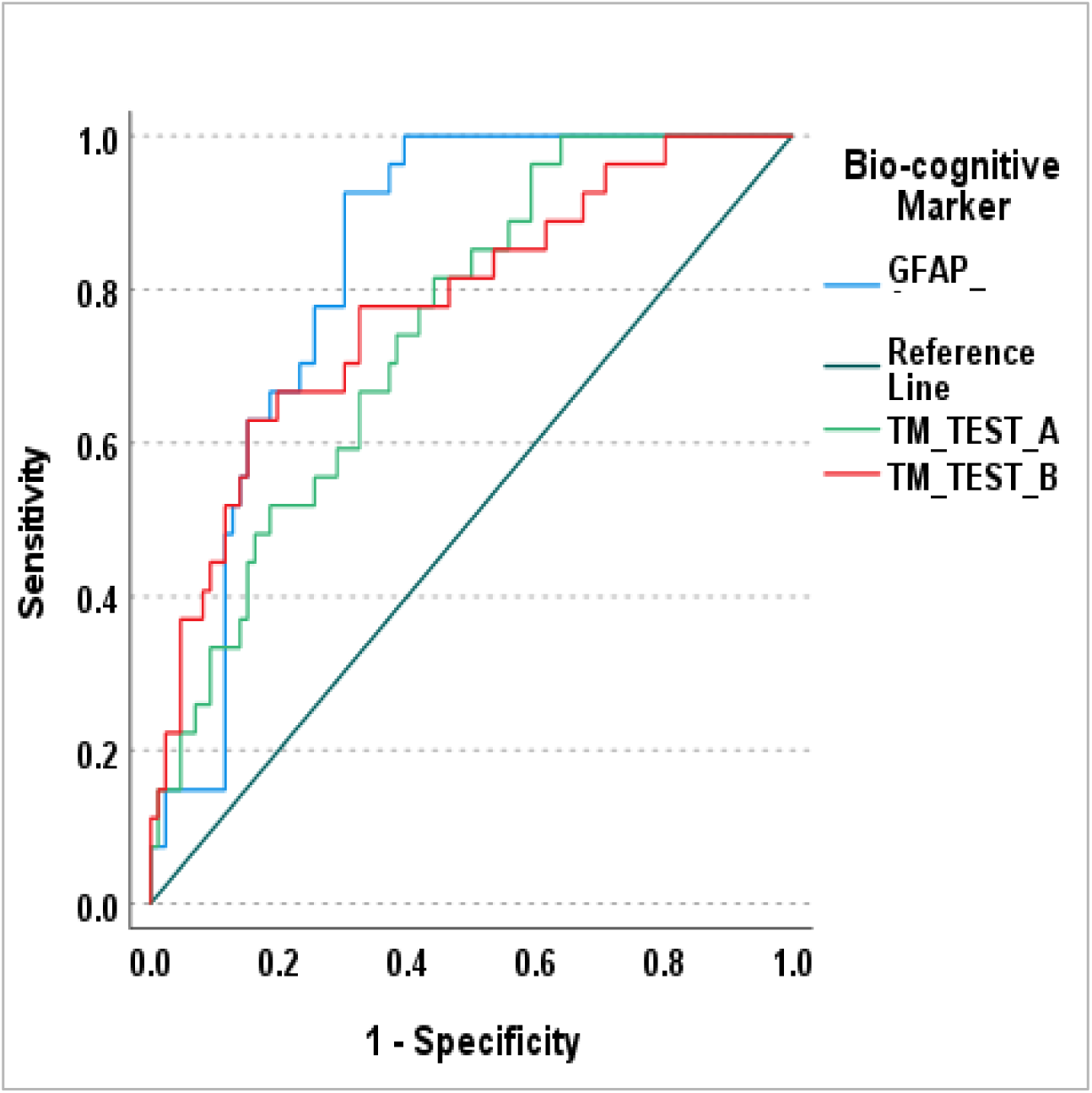
ROC for PD Risk with GFAP & TMT-A/B.

**Figure 3.**
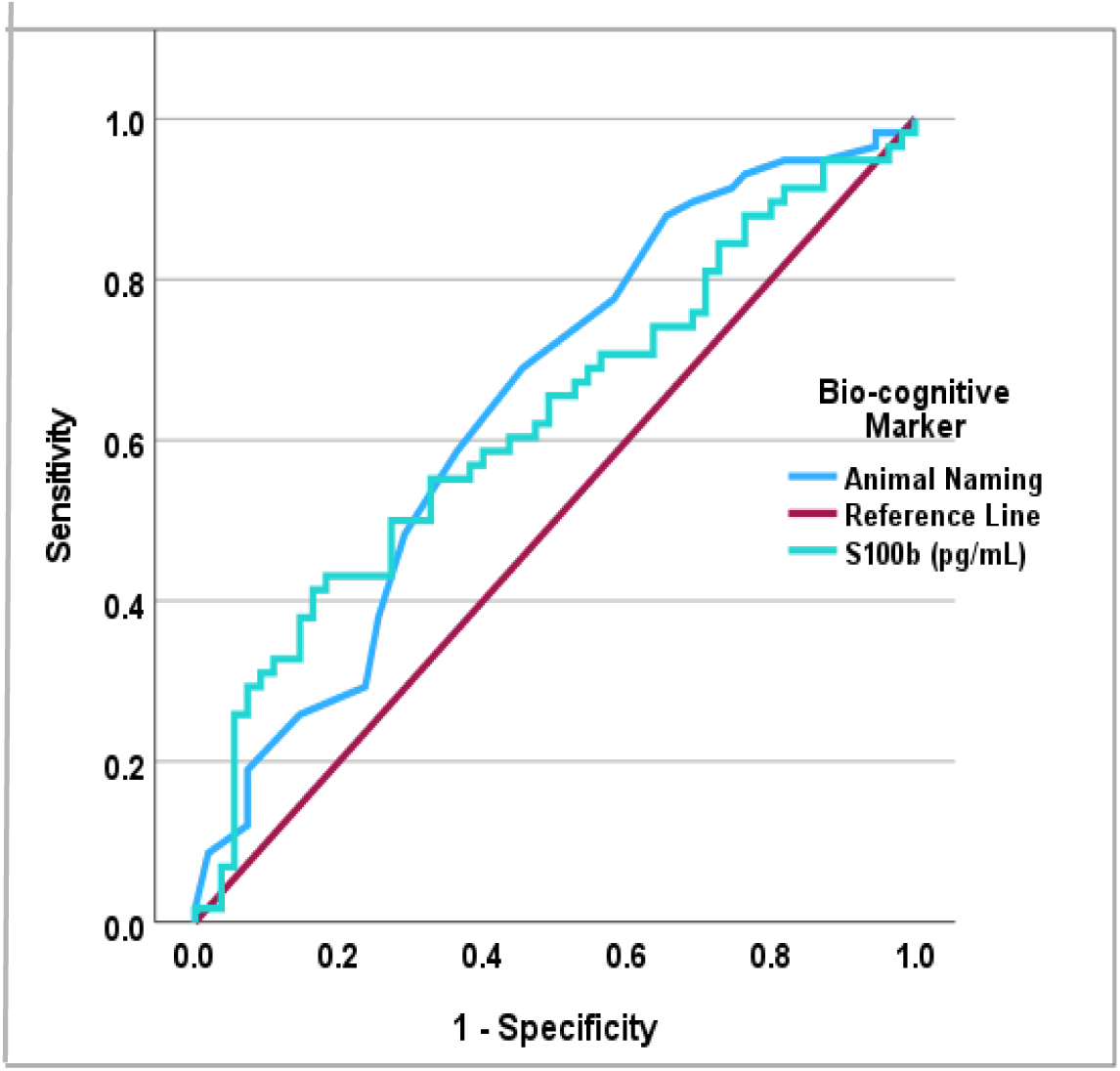
ROC for PD No Risk with S100B & Animal Naming.

**Table 3.** Area Under the Curve for Bio-cognitive Markers Identifying the YES Risk for PD.

| **Table 3.** |  |  |  |  |  |
| --- | --- | --- | --- | --- | --- |
| ***Bio-cognitive markers*** | ***Area*** | ***Std. Error*** | ***Asymptotic Sig*** | ***Asymptotic 95% C. I.*** |  |
|  |  |  |  | ***Lower Bound*** | ***Upper Bound*** |
| GFAP | 0.83 | 0.04 | **0.000** | 0.76 | 0.91 |
| TMT-A | 0.75 | 0.05 | **0.000** | 0.65 | 0.84 |
| TMT-B | 0.81 | 0.04 | **0.000** | 0.73 | 0.89 |
| S100B | 0.63 | 0.05 | **0.018** | 0.53 | 0.73 |
| Animal Naming | 0.65 | 0.05 | **0.005** | 0.57 | 0.77 |

The standardized normative mean for TMT-A in a healthy population with ≥12-years of education in 25 – 34-year-olds is **24.4 (8.71**) which coincides with 93% sensitivity and 60% specificity when entered into the ROC/AUC equation. For TMT-A, Youden’s cut-off is **> 26.98** with a K-S of .40, 81% sensitivity and 45% specificity in ROC coordinates. We found 74% (20/27) of veterans + mTBI outcomes for TMT-A ranged from **26.5 - 58.5**, 30% (9/30) of veteran controls had ranged of **26.7 - 39.7**, and 43% (13/30) of healthy controls had a range of **28.4 - 45.5**. Norms for 67.2 – 72.1-year-old healthy controls have a standardized mean range of **33.8 (6.7) - 40.1 (14.5**). Here, 26.2% (7/27) of subjects with early-stage PD had a range of **43.9-64.8** which is considerably higher than the standardized normative scores for their age and education level.

For the TMT-B, Youden’s cut-off is **>54.3** with a K-S of .35 reflecting 78% sensitivity and 43% specificity. The standardized norm for TMT-B in a healthy population mean age range of 25-34 years old with ≥12-years of education is **50.7 (12.4)** with ROC coordinates of 81% sensitivity and 54% specificity. The results show 67% (18/27) of veterans + mTBI performed within the range of **66.5 - 164.4** which is substantially higher reflecting much worse cognitive flexibility than the standardized norm for their age and education level with 44% (12/27) scoring significantly higher than Youden’s cut-off. There were 56.7% (17/30) of veteran controls that performed within the range of **50.0 - 69.2**, and 50% (15/30) of healthy controls performed within the range of **51.8-81.5**. In the PD group, 67-72-year-olds with ≥12- years of education the TMT-B range is **67.1 (9.3) - 86.3 (24.1).** There were 44% (12/27) PD subjects performed within a range of **88.8-281.6** indicating a sizable impairment in cognitive flexibility and strategic decision-making according to the standardized norms. Taken together, these data show that roughly half of veteran and healthy controls also displayed some decline in cognitive flexibility and decision-making which may be described as premature cognitive aging. This is especially true of veterans + mTBI as performance aligned with early-stage PD senior subjects and was even worse than the standardized norm for senior populations.

The ROC curve for S100B paired with Animal Naming produced an AUC of **.63** and **.67 which** reflected moderate to moderately fair diagnostic accuracy and power as sensitivity and specificity were approximately equal. The cut-off for S100B at >**581.2** *pg/mL* had **74%** sensitivity and **66%** specificity which might be useful for screening rather diagnostic purposes. Of those who had a S100B level *higher than* **581.2** *pg/mL*, 30% (8/27) were veterans + mTBI, 30% were veteran controls (8/27), and 33% (10/30) were healthy controls. More precisely, cut-offs of ≥**695.1** *pg/mL* have **81%** sensitivity and **71%** specificity indicating a **moderate risk** for PD/oND, while a cut-off range of **921.7** to **1159.3** *pg/mL* has **91-95**% sensitivity and **90**% specificity indicate a **strong risk** for PD/oND.

For Animal Naming, the more animals named in 60-seconds, the *<u>better</u>* the categorical and semantic memory. In the age range of 16 – 59 years-old with ≥13-years education, the standardized normative mean score is **21.9 +/- -5.4** which reflects 76% sensitivity and 52% specificity. Youden’s cut-off was **24.0** with 55% sensitivity and 35% specificity. Thirty percent of veterans + mTBI (8/27), 33% of veteran controls (10/30), 63% of healthy controls (19/30), and 70% of PD (19/27) scored ≥24 associated within a 75-90 standardized percentile which indicates negligible categorical or semantic memory problems. Taken together, the S100B + Animal Naming bio-cognitive profile may be useful as a precautionary screening tool to discern the percent of potential risk vs. no risk for PD/oND if monitored over time.

## Discussion

A key issue with blood-based biomarkers is that there is little agreement on metrics to capture a reference range of cut-points with enough sensitivity and specificity to reliably predict disease risks. Case in point, the Food and Drug Administration (FDA) recently announced that the Alzheimer’s plasma biomarker phosphorylated tau 217 (p-tau 217) and β-amyloid 42 (AB42) cut-points approved to detect amyloid neuropathology in 2025 can produce false positives and false negatives and should not be used as a standalone test for diagnostic purposes (Algeciras-Schimnich et al, 2026; ). This finding represents just one of the many debates, challenges, and inconsistencies in determining clinical reference ranges or reliable inter-lab/assay cutoffs that can vary with specimen matrix (serum vs plasma), assay/platform performance, and precision differences (Bago Rozankovic and Simic, 2026; Bazarian et al., 2025). Entrenched scientific dogma that mTBI symptoms do not last longer than 3-months and (5) post-mTBI residual symptoms occur in only 15% of the population is highly debated although growing evidence shows up to 58% of post-mTBI adults experience chronic cognitive impairment lasting longer than 12 months resulting in significant residual effects (Hier et al; McInnes et al., 2020).

Regarding residual effects, remote (∼12-yrs.) mTBI in early middle-age veterans were found to have higher plasma GFAP levels that coincided with worse cognitive flexibility (i.e. TMT-A+B) that unfortunately disappeared in post hoc analyses (Dark et al, 2026). One reason why the relationship between GFAP and cognition was not upheld could be the investigators used log transformations that can inflate or change the direction of outliers (West, 2022). Another TBI biomarker study examining plasma derived exosomes in elder veterans (Ⱦ = 79 yrs ) who experienced a TBI a3- to 5-decades earlier showed GFAP was only minimally helpful in differentiating cognitively impaired (N=35) from unimpaired (N=30) groups (Peltz et al (2020). However, examining exosomes rather than plasma or serum introduces more matrix variability as assays have wide variance in purity, yield, and many labs lack the capability to use this method in addition to lack of assay standardization. Moreover, the Peltz et al., (2020) study allowed different TBI severities and raw cognitive scores for verbal learning and delayed recall were transformed into composite scores. These limitations prevent finding reference ranges and cut-points for predictability. Rather than composite scores, we used raw data of each individual subject to pinpoint those 46% of veterans + mTBI (13/27), 26% of veteran controls (7/27), and 30% of HC (9/30) who had higher levels of GFAP (>39.83 *pg/mL*) that were comparable to 93% of our early-stage PD subjects (25/27).

As noted in our sample, it is important to acknowledge that not everyone experiencing a mTBI will be vulnerable to developing PD since this disease is highly heterogeneous triggered by a number of different factors. Conversely, those who never experience mTBI may encounter myriad factors that indeed put them at risk for PD / oND. Using our reference range cut-points, ∼35% of the combined young cohort (29/84) appear to have an increased risk of developing PD / oND at some point during their lifetime The potential increased risk of PD / oND in our young cohort is somewhat concerning as current world standard population estimates for 30–39-year-olds have ∼29% prevalence estimate (data not available in the United States; Tezen et al., 2026; Nebizadeh et al., 2024). While risk may not equate to prevalence; the fact that ∼35% of the young adults in our sample showed subtle impairments in particular domains of executive function suggests they may be experiencing unrecognized premature cognitive decline portending to advanced aging. Thus, young adults especially young veterans + mTBI showed bio-cognitive aging and appear to be cognitively functioning as if they were late middle-aged to senior adults, early-stage PD, or possibly young onset prodromal PD as there were no observable motor symptoms.

Our bio-cognitive data are supported by Shahim and colleagues (2020) who used magnetic resonance imaging (MRI) and diffusion tensor imaging (DTI) to visually characterize neural aspects of serum GFAP and UCH-L1 levels in mild, moderate, and severe TBI patients after 30-, 90-, 180-days, 1-, 2-, 3-, 4-, and 5-years post-injury (Shahim et al, 2020). These investigators found elevated GFAP first peaked at 3-years that slightly lowered at the 4^th^ and 5^th^ year but *<u>never</u>* returned to baseline levels obtained at 30-days post-injury. At the 3^rd^, 4^th^, and 5^th^ year timepoints, increased GFAP levels were significantly associated with decreased corpus callosum (CC) volume in anterior, mid-anterior, central, and posterior segments. These brain areas are associated with complex sensory integration and specific domains of executive function influencing accuracy in cognitive flexibility, attention, processing speed, and verbal fluency (Barbaresi et al., 2024; Huang et al., 2015). Furthermore, GFAP had stronger diagnostic power ranging from 0.60 to 0.89 during the 4^th^ and 5^th^ years (Shahim et al., 2020) which is similar to our findings of GFAP diagnostic accuracy AUC of 0.83 in veterans +mTBI. A reference range of GFAP levels with clinically relevant cut-points suitable for diagnostic purposes was not reported by Shahim et al. as the median range widely varied from ∼25.0 to 300.0 *pg/dL* which limits clinical utility.

Our data suggests a reference cut-off range was associated with a robust bio-cognitive profile of GFAP+TMT-A+TMT-B where the cognitive outcomes paired with GFAP strengthened the diagnostic accuracy with enough power to predict a risk of PD/oND using a simple blood test. We also identified a cut-point range for S100B+Animal Naming as a potential bio-cognitive screening tool. We were able to determine specific bio-cognitive reference range cut-points and identified the percentage of individuals from every experimental group who *<u>were</u>* and those who *<u>were not</u>* at-risk for PD/oND.

Although serum UCH-L1 received Food and Drug Administration approval in 2018 to determine the need for CT scans in patients admitted into emergency hospitalizations with mild TBI, we found no utility for UCH-L1 in remote mTBI. Again, our results are supported by others who have reported that UCH-L1 does not significantly differentiate among mTBI, moderate TBI, and healthy controls and does not correlate with cognitive function (Shahim et al., 2020). Accordingly, as a housekeeping enzyme important for clearing aggregated protein accumulation in urgent care events, UCH-L1 may be better suited for identifying acute TBI in an ER setting with higher severity levels than mild.

### Limitations and Strengths

To our knowledge, this is the first age- and IQ-match-control study to provide a reference range of cut-points using a bio-cognitive signature in young veterans + mTBI, veteran and healthy controls, and senior early-stage PD subjects to explore if a post-mTBI might be a risk for PD/oND. While our results are promising, they must be viewed with caution as our matched-control proof-of-concept study was a cross-sectional design with relatively small, and unequal group sample sizes. However, conducting this study was critically important for several reasons as we were able to: (1) recognize the subtle, residual effects of ∼5.5-years post-mTBI revealing similar bio-cognitive outcomes in young veterans + mTBI as found in seniors with early-stage PD; (2) quantify laboratory reference range cut-points that accurately defined GFAP+TMT-A+TMT-B as a bio-cognitive signature from which we could discern risks and use against S100B+Animal Naming as a bio-cognitive marker to screen out no-risk individuals; (3) identify potential cut-off values for differentiating prodromal ‘risk’ vs. ‘no risk’ for PD/oND among healthy young controls and those with lifetime mTBI events. Our data will be used to guide future longitudinal research to refine clinically relevant bio-cognitive cut-points to use as markers of disease risks for PD across multiple experimental groups including young healthy controls. Finally, we found BDNF and UCHL-1 were imprecise markers of remote mTBI with no significant relationships with cognitive flexibility, decision-making, or verbal semantic fluency.

### Conclusion

Although the FDA and National Institutes of Health joined to establish TBI biomarker guidelines, (https://www.accessdata.fda.gov/cdrh_docs/reviews/DEN170045.pdf) clinically relevant reference ranges and cut-offs of mTBI-related risks for PD/oND have not been established. Our study is unique as we identified preliminary reference ranges and cut-points to help predict risks vs. no risk for PD/oND. Our results suggest clinicians need to screen young adult patients who have experienced a remote mTBI for subtle decline in cognitive flexibility, decision-making, attention, and semantic verbal fluency in combination with serum GFAP and S100B. A longitudinal follow-up assessment study in this sample is currently underway to examine if these bio-cognitive signatures have prognostic reliability in refining our reference ranges and cut-offs to predict risks for PD/oND. Further, a larger sample size in each of our experimental groups should help validate these findings.

## Data Availability

Only de-identified data produced in the present study will be made available only when Institutional Review Board written approval by the institution requesting the data is confirmed by the corresponding author (VAN).

